# Missense but mis-spliced: germline *TP53* variant c.671A>C (p.E224A) and the path from uncertainty to pathogenicity

**DOI:** 10.1101/2025.07.31.25332437

**Authors:** Irena Velkova, Serena Cappato, Daniela Rivera, Ferruccio Romano, Deborah Schonneger, Renata Bocciardi, Pierre Hainaut, Patrizia De Marco, Viviana Gismondi, Gabriella Cirmena, Ludovica Menta, Marzia Ognibene, Alberto Garaventa, Carla Manzitti, Samuele Brugnara, Yari Ciribilli, Alessandra Bisio, Elisa Marcaccini, Paolo Malatesta, Francesca Faravelli, Paola Menichini, Paola Monti, Valeria Capra

## Abstract

The *TP53* gene encodes the well-known P53 tumor suppressor protein, which plays a crucial role in preventing cancer development. Germline *TP53* variants cause Li-Fraumeni Syndrome (LFS), an autosomal dominant disorder associated with early-onset cancers, including breast cancer, brain tumors, leukemias, bone cancers, and soft tissue sarcomas. Functional studies in yeast and human cells demonstrated that *TP53* variants can have various effects, such as partial or complete loss of function and even gain of pro-oncogenic activities.

Here, we identified a germline *TP53* variant c.671A>C, resulting in the missense mutant protein p.E224A in the context of early-onset retroperitoneal rhabdomyosarcoma occurring in a child with a notable family history of cancer, suggestive of LFS. The variant was initially classified as a variant of uncertain significance (VUS). Functional assays in yeast and human cells demonstrated wild type-like activity of the protein p.E224A; however, *in silico* analysis predicted at RNA level a splicing defect, which we further investigated using a minigene approach. This analysis showed that the variant c.671A>C causes the skipping of exon 6, potentially introducing a frameshift in cDNA and a premature stop codon, which likely triggers nonsense-mediated mRNA decay; the loss of heterozygosity at the c.671 position in the parent’s *TP53* transcript further confirmed the splicing impairment.

In summary, these findings supported reclassifying the *TP53* germline variant c.671A>C (p.E224A) from VUS to likely pathogenic, providing a definitive molecular diagnosis for family counseling. Additionally, this study sheds light on how certain *TP53* variants that are defined as missense, can be linked to disease mechanisms through RNA splicing disruption, highlighting the need for their deep functional assessment.

## Introduction

The human *TP53* gene, located on the short arm of chromosome 17 (17p13.1), encodes the well-known P53 tumor suppressor protein, which was originally identified as a host protein bound to the SV40 large T antigen in virally transformed cells [1,2]. P53, commonly referred to as the guardian of the genome [3,4] functions primarily in the cells as tetrameric transcription factor that binds specific DNA sequences to regulate the expression of a plethora of downstream effector genes; these genes play a critical role in key cellular processes such as cell cycle arrest, DNA repair, apoptosis and senescence, thereby stopping the proliferation of damaged cells, which is a critical step in cancer prevention. More recently, P53 has been shown to have a broader range of functions that enhance its anti-cancer activity; indeed, emerging research reveals its involvement in metabolic regulation, ferroptosis, and immune system modulation, further underscoring the pivotal role of P53 in maintaining cellular homeostasis and preventing the initiation and progression of cancer [4,5].

Somatic *TP53* variants occur in a wide range of tumor types, and when in the germline, they cause the development of Li-Fraumeni Syndrome (LFS), an autosomal dominant genetic disorder that significantly increases the risk of developing multiple cancers [6,7]. The diagnosis of LFS traditionally relies on Chompret’s criteria, an evolving set of clinical classification guidelines that include a family history of cancer, multiple primary tumors in one individual, early onset of breast cancer, the occurrence of rare tumor types, and cancers affecting children and adolescents [8]. Specifically, the spectrum of cancers seen in LFS patients includes soft tissue sarcomas, osteosarcomas, breast cancer, brain tumors, leukemia, and adrenocortical carcinoma. These cancers typically develop at an earlier age compared to the general population, underscoring the critical need for early detection and surveillance in LFS [8]. Among *TP53* alterations identified in LFS patients, missense variants are the most common, accounting for approximately 80% of cases; they are mainly located in the central DNA binding domain of the P53 protein, with eights hotspot mutations (*e.g*., p.R175H, p.G245D, p.G245S, p.R248Q, p.R248W, p.R273C, p.R273H and p.R282W) associated with a poorer prognosis.

Besides missense mutations, other *TP53* alterations reported in LFS patients include frameshift, nonsense, and splicing variants, which can disrupt the normal function of the P53 protein in different ways, further contributing to increasing cancer risk in LFS affected individuals (https://tp53.isb-cgc.org/).

Both small- and large-scale systematic studies have been conducted to assess the impact of *TP53* variants on the biological functions of the P53 protein, often using yeast models because of their simplicity and versatility for studying protein function [9–13]. One of the key early studies by Kato et colleagues evaluated 2,134 missense *TP53* variants and their effects on the protein’s transactivation activity, revealing significant functional heterogeneity among mutant P53 proteins. This study demonstrated that *TP53* variants can be classified into several categories based on their impact on protein function, such as loss of function, partial function, wild type-like, super-transactivating, and those with altered specificity [14]. Furthermore, certain *TP53* variants can acquire pro-oncogenic properties through dominant negative effects (*i.e.*, interference with the function of the wild type counterpart when both are expressed in the same cell) or through neo-morphic activities. This latter mechanism occurs when the mutant P53 protein gains new functions that promote cancer (*i.e.*, gain of function, GOF); as an example, mutant P53s can interact with other proteins, such as members of the P53 family (*i.e.*, P63 and P73), leading to aberrant protein aggregation and enhanced oncogenic potential [15].

Since yeast models lack the full complexity of the P53 regulatory network, additional functional screens have been conducted in human cells [16–18]. More recently, a large-scale study was conducted by introducing 9,225 *TP53* variants into cancer cells harboring a wild type *TP53* locus, using the CRISPR-based technology. Unlike previous cDNA overexpression screens, this approach maintains the physiological regulation of the *TP53* gene, including the function of endogenous promoters, enhancers, alternative splicing, and microRNA binding sites [19]. By preserving these natural regulatory mechanisms, this study offers a more accurate representation of how *TP53* variants might function, providing deeper insights into their biological effects.

The increasing knowledge about *TP53* variants and their diverse effects on the P53 protein is crucial for advancing integrated genotypic and phenotypic analyses in LFS. This integration is necessary for enhancing cancer risk prediction and developing personalized more effective strategies for higher risk LFS individuals [20–28]. However, a significant challenge in implementing such personalized approaches is the rarity of many individual *TP53* variants, which complicates the effort to precisely understand their role and the associated molecular mechanism driving cancer development.

Here, we described comprehensive functional studies we have undertaken on a novel heterozygous germline *TP53* variant (c.671A>C, p.E224A) identified in a non-consanguineous family having an aggregated history of typical LFS cancers.

## Materials and Methods

### WES and Sanger analysis

Genomic DNA from peripheral whole blood of the proband was extracted using QIAsymphony DNA Kit (Qiagen, Hilden, Germany) and quantified using NanoDrop 2000c Spectrophotometer and Qubit dsDNA HS and BR Assay Kit (ThermoFisher Scientific, Carlsbad, CA, USA). For the exome library preparation, 200 ng of genomic DNA was used with the xGen® Exome Research Panel v2.0 targets (Integrated DNA Technologies, IDT, Coralville, Iowa), consisting of 415,115 probes (approximately 19,435 genes) that span a 34 Mb target region of the human genome and 39 Mb of probe space. Quantified DNA library was loaded on a flow cell for subsequent cluster generation. The sample was paired-end sequenced on an Illumina Novaseq 6000 platform (Illumina, San Diego, CA, USA) with a read length of 2×151 bp to achieve a minimum of 150× on-target coverage. The FASTQ files were aligned to the hg38 human reference genome using the Burrows-Wheeler Aligner package version 6.1. Variant calling was performed with Genome Analysis Tool Kit (GATK4) and variants were annotated with Annovar (database updated May 27, 2019) [29]. We prioritized variants based on i) allelic frequency (MAF) minor or equal to 0.01 in GnomAD v2.1.1, ii) prediction as deleterious or damaging by PolyPhen and SIFT and iii) with a CADD value of at least 20. The *TP53* variant identified by WES was confirmed by amplification of exon 6 with the Platinum PCR SuperMix High Fidelity kit (Invitrogen, ThermoFisher Scientific, USA), using the following forward 5’-GTGAGCAGCTGGGGCTGG-3’ and reverse 5’-GAGGTCAAATAAGCAGCAGG-3’ primers. The PCR product was purified using the GenUP Exo SAP kit (BiotechRabbit, Berlin, Germany) and sequenced bidirectionally using the Big Dye Terminator Cycle Sequencing kit (Applied Biosystems, Foster City, CA, USA) on a 3500XL platform (Applied Biosystems).

### Variant interpretation

Publicly available algorithms that predict functional consequences of genetic variants were used (*i.e.*, Align-GVGD, A-GVGD; Combined Annotation Dependent Depletion, CADD; Polymorphism Phenotyping v2, PolyPhen2; Sorting Intolerant From Tolerant, SIFT; MutationTaster; Rare Exome Variant Ensemble Learner, REVEL; Multivariate Analysis of Protein Polymorphism, MAPP and BayesDel). Variants functional effect on splicing was evaluated using MaxEntScan (MES), SpliceSiteFinder (SSF), GeneSplicer (NNSPLICE), and SpliceAI; the dbscSNV Ada and dbscSNV RF databases of pre-computed prediction scores were also interrogated. The ClinVar and LOVD status, along with a literature review were performed. The variant was classified according to ClinGen *TP53* Expert Panel Specifications to the ACMG/AMP Variant Interpretation Guidelines for *TP53* (Version 2.2.0 released in September 2024) [30].

### Yeast functional assays

Yeast isogenic reporter strains (yLFM-P21-5’, yLFM-MDM2P2C, yLFM-BAXA+B and yLFM-PUMA) were used [31]. Yeast cells were grown in YPDA medium (1% yeast extract, 2% peptone, 2% dextrose, 200 mg/L adenine) or selective medium (w/o 2% agar), containing dextrose or raffinose as a carbon source plus adenine (200 mg/L); leucine and/or tryptophan were added as selection markers (Sigma-Aldrich, Saint Louis, Missouri, USA). Galactose was added to the medium to modulate P53 expression. Construction of yeast expression vectors and reporter assays along with the corresponding western blot procedures are detailed in the <u>Supplementary Material.</u>

The transactivation and the dominant negative ability of mutant P53 proteins were calculated as percentage with respect to wild type P53 protein (100%); data were presented as fold induction over empty vector(s) (pRS314; pRS314 plus pRS315) ± standard deviation. We previously classified a mutant P53 protein as Loss Of Function (*i.e.*, LOF<25%) or Partial Function (*i.e.*, PF≥25%) [21]; in the present paper, we also defined a mutant P53 protein as wild type-like (i.e., ≥80%). Regarding the dominant negative ability, a mutant P53 protein was previously classified as dominant negative (DN) when the activity was <90%, otherwise recessive (REC) (i.e., ≥90%) [21]; here, due to the different experimental set-up, we adopted the cut-off <75% and ≥75% to define DN and REC mutant P53 proteins, respectively.

### Mammalian functional assays

The human colon carcinoma cell line HCT116*^TP53^*^-/-^ (Dr. B. Vogelstein, the Johns Hopkins Kimmel Cancer Center, Baltimore, MD) were cultured in Roswell Park Memorial Institute 1640 medium (RPMI, Euroclone, Milan, Italy) supplemented with 10% fetal bovine serum (FBS, Euroclone), 1% L-Glutamine (Euroclone) and 1% Penicillin/Streptomycin (Euroclone). Construction of mammalian expression vectors and reporter assays along with the corresponding western blot procedures are detailed in the <u>Supplementary Material</u>. The functional classification of mutant P53 proteins was based on the cut-off indicated above; data were presented as fold induction over empty vector (pCI-neo) ± standard deviation.

### Colony formation assay

HCT116*^TP53^*^-/-^ were seeded into 24-well plates at 1.50 x 10^5^ cells/well and incubated for 24 hours at 37°C. On the next day, cells were transfected with 200 ng of vector expressing P53 protein (wild type and mutant) or with the empty vector (pCI-neo) in the presence of Mirus Bio^TM^ TransIT^TM^-LT1 Transfection Reagent (Mirus Bio, Madison, WI, USA). After 24 hours, cells were collected, counted with the TC20 cell counter (Bio-Rad, Milan, Italy), and seeded at a density of 300 cells/well in 6-well plates in the presence of G418 selection (Roche, Basel, Switzerland). After 12 days of incubation, the colonies were directly stained with 1% methylene blue for 30 minutes, washed with tap water, and counted under a microscope. The average number was calculated for each experimental condition, considering only colonies containing more than 40 cells. Data are shown as the mean number of colonies per well ± standard deviation.

### Minigene splicing assay

To perform the functional analysis of the c.671A>C *TP53* variant by the minigene approach, the fragment spanning the *TP53* exon 6 (GRCh38/hg38, chr17:7,674,754-7,675,052) was amplified by PCR from genomic DNA of the proband’s parent. The mutant *TP53* allele, along with wild type, was subcloned into the EcoRI site of the splicing vector pSPL3 [32] and checked by Sanger sequencing. The human embryonic kidney HEK293 cell line (ATCC) was first used and cultured in Essential Modified Medium (MEM) with 10% FBS. Cells were seeded at 5×10^5^/well into 6-well plates, incubated for 24 hours at 37°C, and transfected with 2 µg of pSPL3-based vectors (empty, wild type and mutant) by Lipofectamine 2000 (ThermoFisher Scientific). After 24 hours, cells were harvested and processed for RNA extraction (RNeasy plus Mini Kit, Qiagen, Germany); 500 ng of total RNA was retrotranscribed with the Advantage® RT-for-PCR (Takara, Shiga, Japan). cDNA was used as template for PCR amplification (GoTaq Master mix, Promega, Madison, WI, USA) with forward and reverse specific oligonucleotides (5’-TCTGAGTCACCTGGACAACC-3’; 5’-ATCTCAGTGGTATTTGTGAGC-3’) for the vector’s exons. Other cell lines, including H1299 (*i.e.*, lung cancer type), HCT116, HCT116*^TP53^*^-/-^, and MCF7 (*i.e.*, breast cancer type), were used to confirm the results in different cellular backgrounds.

### Analysis of *TP53* expression in the proband’s parent

Peripheral blood sample was collected in PAXgene Blood RNA Tubes (Qiagen) from the proband’s parent carrying the *TP53* variant; total RNA was extracted with the PAXgene Blood RNA Kit (Qiagen) and converted into cDNA as described above. PCR amplification on cDNA was subjected to Sanger sequencing with P3, P4, P5 and P6 primers [33,34].

### Analysis of *TP53* variants recurrence at codon 224 in somatic and germline datasets

The frequencies of missense *TP53* variants at codon 224 were retrieved from NCI *TP53* database (https://tp53.isb-cgc.org/; corresponding to the R20 version of the IARC TP53 database for germline variants), COSMIC database (somatic variants in cancer; downloaded on November 12, 2024) and Genome Aggregation Database (gnomAD, v. 4.1.0). To predict the effects of the *TP53* variants on splicing, SpliceAI algorithm was used.

### Analysis of RNA levels and splicing patterns of *TP53* variants at codon 224

Data on mRNA levels for *TP53* variants at codon 224, compared to wild type expression, were retrieved from public RNA-Seq datasets including TCGA (The Cancer Genome Atlas Program), comprising 33 cancer cohorts extracted and combined from firebrowse (http://firebrowse.org) and CCLE (Cancer Cell Line Encyclopedia, DepMap Public 24Q2 release). For TCGA, read counts were normalized across the combined dataset using the DESeq2 package (v1.46.0), excluding genes with less than 10 total read counts across all samples. Levels of mRNA expression for each patient were matched to their variant profiles, retrieved from cBioPortal (https://www.cbioportal.org/). Splicing patterns were further examined using raw RNA-Seq data (available through NCI, dbGaP Study Accession: phs000178.v11.p8) and visualized with Sashimi plots using the ggsashimi tool [35]. For CCLE, mRNA levels in cell lines carrying homozygous codon 224 *TP53* variants were retrieved through DepMap (https://depmap.org/portal/data_page/?tab=allData); RNA levels were statistically compared to wild type cell lines using the Wilcoxon rank-sum test in R, and visualized using the ggplot2 package.

### Statistical analysis

One-way and two-way ANOVA with multiple comparisons was performed using GraphPad Prism (version 9.4.0, GraphPad Software, San Diego, CA, USA); statistical significance is reported in each figure. A *p*<0.05 was considered statistically significant.

## Results

### Identification of the germline *TP53* variant c.671A>C (p.E224A)

WES analysis was performed on the constitutive DNA of the proband, identifying the variant c.671A>C (p.E224A) in the *TP53* gene (NM_000546.5; NP_000537.3). Segregation analysis along with Sanger sequencing demonstrated that it was inherited through the maternal lineage, leading to a diagnosis of LFS syndrome for the proband due to the oncological phenotype of the family. WES also identified two other variants: c.6337G>A (p.V2113M) in the *RYR2* gene and c.280G>A (p.D94N) in the *GNAI2* gene. Mutations in the *RYR2* gene, encoding a ryanodine receptor found in the sarcoplasmic reticulum of the cardiac muscle, are associated with rare, potentially lethal inherited arrhythmia disorders, while mutations in *GNAI2* gene, encoding an α subunit of guanine nucleotide binding proteins (G proteins), cause developmental abnormalities and immune dysregulation; these variants are probably linked to the Paroxysmal Supraventricular Tachycardia (PSVT) episodes occurred during the first admission of the proband.

The *TP53* germline variant c.671A>C is reported in ClinVar as of uncertain significance, as well as unknown/not classified in LOVD (**Supplementary Table 1**). The variant, not present in population databases, is located in exon 6 of the *TP53* gene, and replaces the moderately conserved glutamic acid in position 224 with an alanine (p.E224A), an amino acid with dissimilar properties. *In silico* functional analyses suggested that the p.E224A amino acid change is likely detrimental to both the structure and function of the P53 protein; additionally, specific bioinformatic tools predicted the loss of the canonical donor splice site (**Supplementary Table 1**).

### The P53 variant p.E224A shows wild type-like transactivation activity and the absence of dominant negative potential in yeast reporter assays

The transactivation activity of the mutant P53 protein p.E224A was evaluated using four isogenic yeast reporter strains (i.e., yFLM-P21-5’, yLFM-MDM2P2C, yLFM-BAX A+B and yLFM-PUMA) in comparison with wild type P53 and the LOF mutant p.R175H; the activity was analyzed by growing cells at 30°C (*i.e.*, physiological yeast growth temperature) and 37°C (*i.e.*, physiological human cells growth temperature) to identify possible temperature-sensitive behaviour of the mutant P53 protein under study.

The variant p.E224A showed a transactivation ability comparable to wild type P53 across all reporter strains at both temperatures, behaving like a wild type-like P53 protein. In contrast, as expected, the LOF p.R175H variant showed no activity (**Figure 1A, Supplementary Table 2).** Western blot analysis confirmed that the observed functional differences between p.E224A and p.R175H were not due to variation in protein expression levels (**Supplementary Figure 1A).**

**Figure 1.**
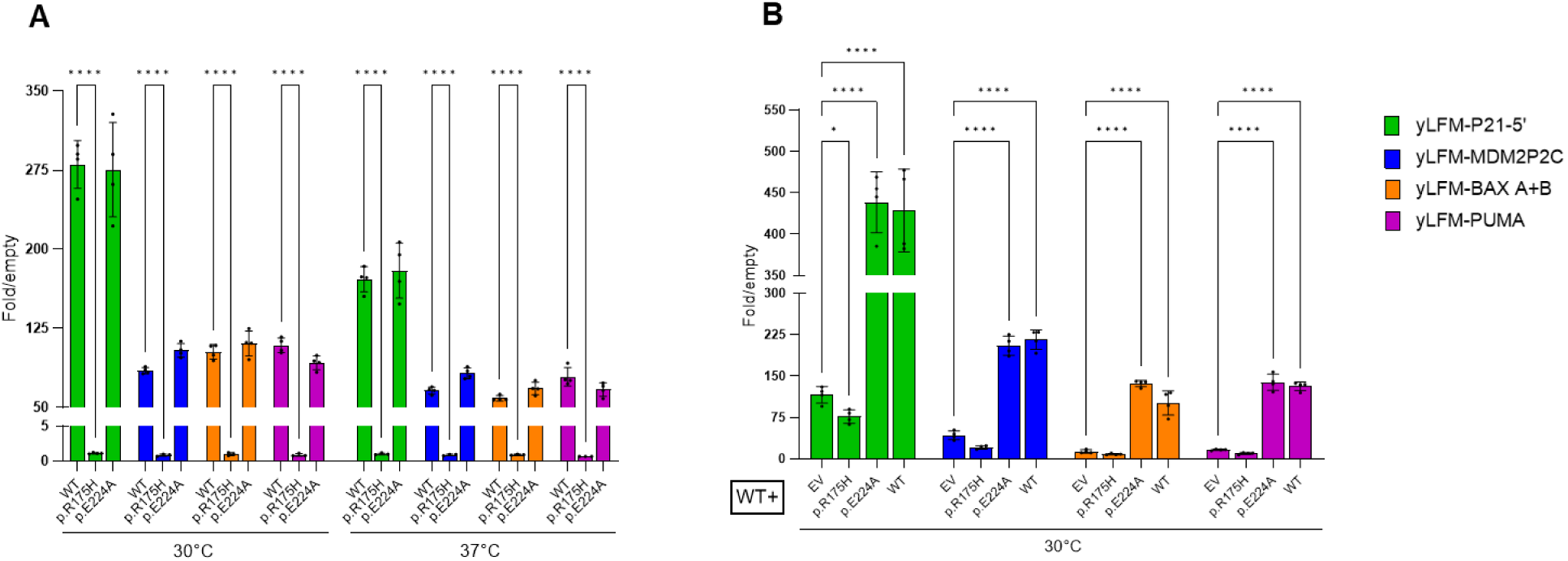
Impact of P53 variant p.E224A on transactivation and dominant negative activity in yFLM-P21-5’, yLFM-MDM2P2C, yLFM-BAX A+B, and yLFM-PUMA yeast reporter strains. The p.R175H variant was introduced in the analysis as a control of LOF and DN P53 protein. A) Transactivation (galactose 0.128%) and B) dominant negative activity (galactose 0.016%) are shown as the average of folds of induction over empty vector(s), derived from at least three measurements. Only significant comparisons are indicated: *p≤0.05, ****p≤0.0001, WT=wild type, EV=empty vector.

Given the heterozygous status of the mutant p.E224A in the affected proband, the dominant negative activity (*i.e.*, inhibition of wild type P53 activity) of the variant was evaluated in the same yeast strains alongside p.R175H; this feature was analyzed exclusively at 30°C, as the activity of both mutant P53 proteins was unaffected by temperature. The variant p.E224A was unable to inhibit the activity of wild type P53 on all reporter strains, acting similarly to the wild-type P53 protein; the p.R175H variant, as expected, showed a dominant negative ability (**Figure 1B; Supplementary Table 2).** Western blot analysis further confirmed that the results were not caused by differences in the expression levels of the mutant P53 proteins (**Supplementary Figure 1B**).

In conclusion, the results of the yeast reporter assays indicated that the variant p.E224A exhibits an activity comparable to that of the wild-type P53 protein.

### The P53 variant p.E224A shows both wild type-like transactivation and clonogenic activity in mammalian cells

The transactivation activity of the mutant P53 protein p.E224A was studied in mammalian HCT116*^TP53^*^-/-^ cells in comparison with wild type P53 and the LOF mutant p.R175H as previously. The variant p.E224A exhibited a transactivation activity comparable to wild type P53 protein across all promoter reporters tested (*i.e.*, pGL3-P21, pGL3-MDM2 and pGL3-BAX), whereas the mutant p.R175H, used as control of LOF P53 protein, confirmed the lack of transactivation ability (**Figure 2A, Supplementary Table 2**). Western blot analysis showed comparable expression levels of all P53 proteins, indicating that the results from the mammalian cells-based reporter assay were also not due to differences in P53 protein expression (**Figure 2B**). Moreover, the analysis of the endogenous P21 protein level, as a target of P53 transcriptional activity, revealed equivalent induction by p.E224A and wild type P53 protein; in contrast, p.R175H failed to induce the P21 protein as observed for the empty vector (**Figure 2B**). Lastly, the colony-forming ability was evaluated in HCT116*^TP53^*^-/-^ cells expressing the P53 variant p.E224A. These cells exhibited clonogenic activity comparable to those expressing wild type P53, whereas cells expressing the variant p.R175H formed the highest number of colonies (**Figure 2C**).

**Figure 2.**
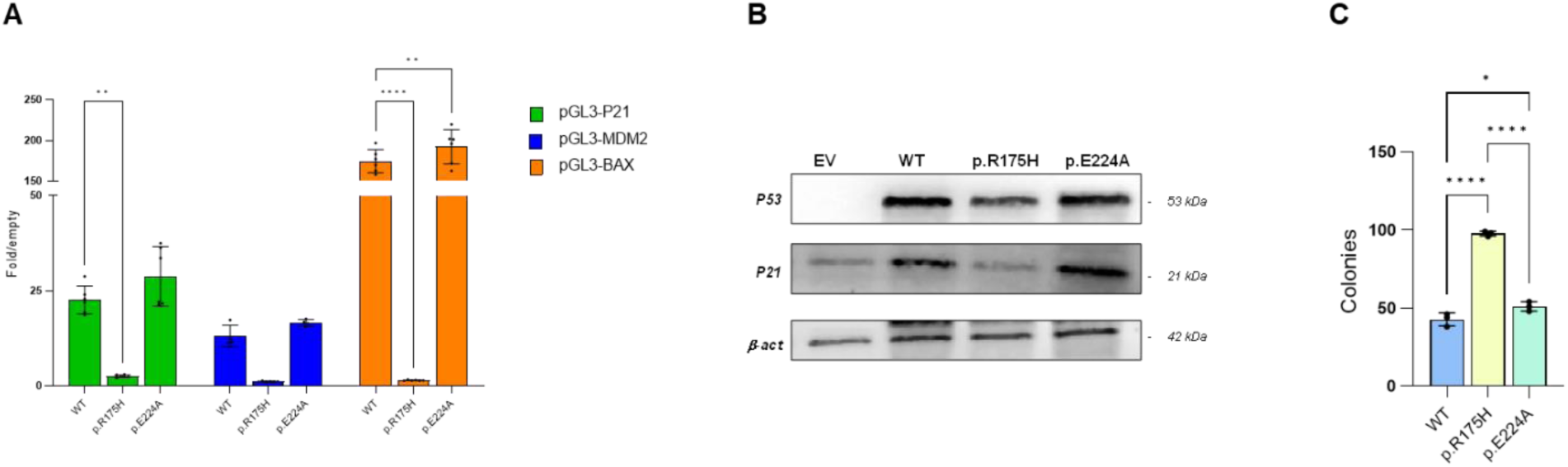
Impact of P53 variant p.E224A on transactivation and clonogenic activity in HCT116^TP53-/-^ cells. The variant p.R175H was introduced in the analysis as a control of LOF P53 protein. A) Transactivation activity on pGL3-P21, pGL3-MDM2, and pGL3-BAX reporters is shown as the average of folds of induction over the empty vector from at least five measurements. B) Representative Western blot showing the level of P53 transfected proteins and endogenous P21; β-actin was used as a loading control. C) Colony forming ability of cells transfected with different P53 proteins expressed as the number of colonies. Only significant comparisons are indicated: *p≤0.05, **p≤0.01, ****p≤0.0001.

Collectively, these results confirm that the P53 variant p.E224A behaves like the wild type P53 protein in mammalian cells, consistent with the observations from the yeast reporter assays.

### The *TP53* variant c.671A>C affects the splicing at exon 6 of *TP53* gene

Based on *in silico* predictions on the *TP53* splicing **(Supplementary Table 1)**, the impact of *TP53* variant c.671A>C was evaluated by a minigene approach, by cloning the genomic fragment from the proband’s parent spanning the *TP53* exon 6 in the pSPL3 splicing vector.

While HEK293 cells transfected with the wild type minigene generated two bands, corresponding to alternative splicing with or without the inclusion of exon 6 (376 bp and 263 bp, respectively), cells transfected with the c.671A>C variant construct expressed only transcripts with the skipping of the *TP53* exon 6 (*i.e.*, 263 bp); as expected, a band of 263 bp was obtained from cells transfected with the empty vector (**Figure 3A**). These results were recapitulated in other cell lines (*i.e.*, H1299, HCT116 with wild-type and null *TP53* status, and MCF7), which are indicative of cancer types developed in the identified LFS family (**Supplementary Figure 2**).

**Figure 3.**
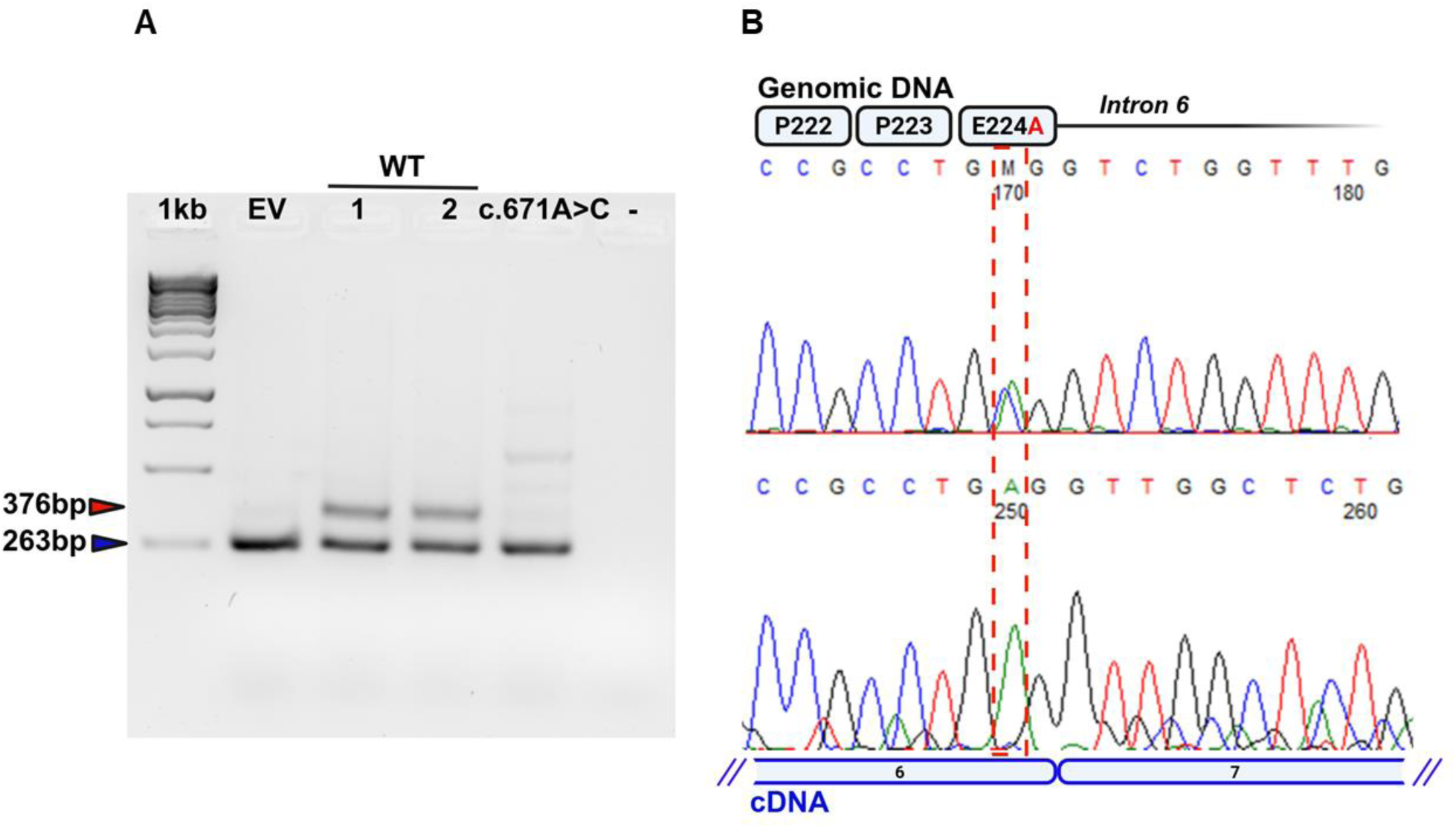
Impact of *TP53* variant c.671A>C on splicing. A) Wild type and mutant genomic fragments spanning exon 6 and the intronic flanking sequences were subcloned into the pSPL3 splicing vector and transfected into the HEK293 cell line. PCR products obtained from empty vector (EV), two different wild type (WT) clones, a single c.671A>C clone, and the PCR negative control without template are shown. B) Sequencing of the PCR products obtained from genomic DNA and RNA (cDNA) of the proband’s parent spanning the exon 6/7 junction. Red dashed lines include the c.671 nucleotide position.

Lastly, the *TP53* mRNA from the whole blood of the proband’s parent was analyzed; a loss of heterozygosity at c.671 nucleotide position of *TP53* mRNA, in which only the wild type allele was detectable (c.671A), was observed compared to genomic DNA (c. 671A/C) (**Figure 3B**).

All these results indicated that the *TP53* variant c.671A>C promotes the complete exclusion of exon 6 from the transcript, likely resulting in a frameshift and the introduction of a premature stop codon. As a result, the aberrant transcript could be targeted for degradation via nonsense-mediated mRNA decay (NMD), thus resulting in the lack of expression of the corresponding P53 protein p.E224A.

### *TP53* variants at codon 224 generally affect splicing with a decrease in RNA expression

To explore the general impact of *TP53* variants at codon 224 of the P53 protein, a total of seven other variants were identified across the germinal, COSMIC, and gnomAD *TP53* datasets; these variants are responsible for aminoacidic substitutions, including one synonymous. While five of these variants have a significant predicted SpliceAI score (>0.7), two variants (*i.e.*, c.670G>A, p.E224K and c.670G>C, p.E224Q), which occur at the first nucleotide position of the codon, show a weak effect on splicing predictions (SpliceAI < 0.1) (**Supplementary Table 3**). Moreover, while the c.672G>T variant encoding for P53 protein p.E224D is the only nucleotide substitution documented in the germline database as well as the most frequent mutation in somatic cancer COSMIC datasets, the c.671A>C (p.E224A) *TP53* variant, we identified in the present study has never been reported in the available public datasets (**Supplementary Table 3**). The mRNA expression levels of *TP53* variants at codon 224, when available, were also analyzed from TCGA and CCLE databases. Two *TP53* variants at codon 224 were documented, comprising the missense p.E224D (c.672G>C or c.672G>T) and the synonymous p.E224E (c.672G>A); for both *TP53* variants, a decrease in RNA expression was observed compared to wild type expressing controls (**Figure 4)**. This decrease was significant for both variants in the TCGA dataset and only for p.E224D in the CCLE dataset, since p.E224E could not be statistically evaluated in the latter dataset (*i.e.*, presence of only one documented cell line). Lastly, RNAseq data from TCGA patients were further inspected for splicing alterations, confirming the overall decreased expression and some retention of intron 6 with respect to normal tissues (**Supplementary Figure 3**). Notably, this pattern was not observed in three patients (*i.e.*, SARC-p.E224D, c.672G>T; OV- and LGG-p.E224E, c.672G>A), retaining normal *TP53* expression levels and splicing patterns. Accordingly, copy number analysis from cBioportal indicated the presence of a wild type *TP53* allele alongside with the mutant, which may reflect *TP53* diploidy, or shallow deletion in tumor cells or a high proportion of non-cancerous cells in the sample.

**Figure 4.**
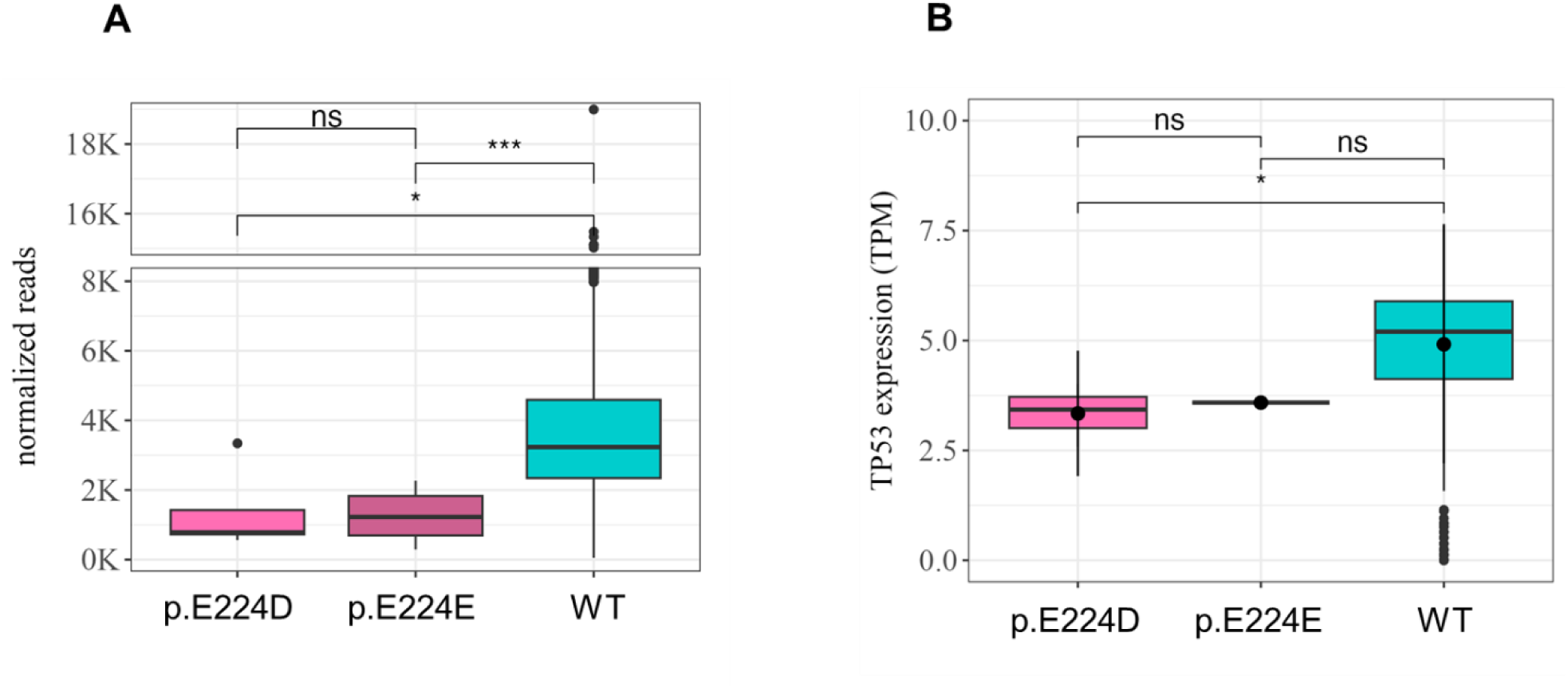
mRNA expression of TP53 variants p.E224D and p.E224E retrieved from public datasets. mRNA expression of A) TCGA patients and B) CCLE cell lines were compared to wild type (WT) expressing controls. The *TP53* variant p.E224D (c.672G>T) is present in the TC106 (ID ACH-001283; Ewing sarcoma), RERFLCKJ (ID ACH-000482, Non-small cell lung cancer) and NCIH716 (ID ACH-000491, Colon Adenocarcinoma) cell lines; the variant p.E224E (c.672G>A) is present in SNU398 (ID ACH-000221, Hepatocellular Carcinoma) cell line. *p≤0.05, ***p≤0.001, ns=not significant, WT=wild type.

Taken together, these results support the hypothesis that *TP53* variants at codon 224, specifically those affecting the second and third nucleotide position, may disrupt the donor splice site at the exon/intron 6 boundary, resulting in a read-through transcript that is subsequently targeted by NMD.

## Discussion

The *TP53* gene encodes the essential tumor suppressor protein P53, and its alterations are commonly associated with cancer development. In particular, pathogenic germline variants are linked to LFS, a cancer predisposition disorder characterized by a wide range of tumors, early onset of cancer, increased sensitivity to genotoxic agents, and a high incidence of multiple tumors. Identifying cancer patients who carry pathogenic or likely pathogenic germline *TP53* variants is crucial for tailoring treatment strategies, optimizing follow-up care, and providing pre-symptomatic testing for at-risk family members [36].

Here, we described a case of a retroperitoneal rhabdomyosarcoma in a child who died from this pathology; the child had a family history of cancer, with a phenotypic spectrum attributable to LFS. WES analysis identified the germline missense *TP53* variant c.671A>C, encoding the p.E224A protein that, to date was never reported in public datasets; to our knowledge, this is the first report of a LFS case with an early-onset embryonal rhabdomyosarcoma carrying this specific *TP53* variant. In general, sarcoma represents a hallmark of LFS, accounting for about one-fourth of tumors in affected individuals, with rhabdomyosarcoma and osteosarcoma being the most common subtypes. The c.671A>C *TP53* variant had been previously identified in one LFS patient with childhood-onset adrenal cortical carcinoma [37] and in one patient with non-small-cell lung cancer (NSCLC), where it co-occurred with ALK rearrangements at the somatic level [38]. Notably, the proband exhibited multiple bilateral suspicious secondary lesions on a CT pulmonary scan. All these evidences supported the hypothesis that the *TP53* variant c.671A>C (p.E224A) might contribute to the clinical phenotype we observed.

However, *in silico* predictions alone were insufficient for the definitive *TP53* variant classification. Indeed, we applied the ClinGen *TP53* Expert Panel Specifications for the ACMG/AMP Variant Interpretation Guidelines (Version 1.2, approved in April 2019 and released in March 2021) [30]; briefly, these specifications adapt the standard ACMG/AMP criteria to the unique characteristics of pathogenic variants in the *TP53* gene. The following classification criteria toward pathogenicity for the *TP53* variant c.671A>C (p.E224A) were applied: PM2 supporting (*i.e.*, absence of the variant from population databases) and PP3 supporting (*i.e.*, A-GVGD: C35; BayesDel: 0.17; *i.e.*, concordant scores to support a deleterious effect of the variant on the gene or gene product) (**Supplementary Table 2**, **version 1.2**). According to ClinGen guidelines, meeting these two criteria was not sufficient to classify the variant, being categorized as a variant of uncertain significance (VUS).

The *TP53* variant c.671A>C (p.E224A) is located in the DNA binding domain of the P53 protein, but it was reported to have functional transactivation in the yeast-based assay developed by Kato and colleagues [13]; in addition, studies conducted in human cell lines confirmed that this alteration is proficient in growth suppression [17,18]. Given the suggestive family history and the patient’s clinical progress, along with *in silico* predictions, we decided to perform deep functional analyses.

We took advantage of our experimental system that exploits yeast reporter strains differing only in the p53 target response element of interest, in an otherwise isogenic background. Our results demonstrated a wild type-like transactivation ability (*i.e*., average activity 103% with respect to wild type) of P53 protein p.E224A; moreover, our yeast dominance assay, which resembles the *TP53* heterozygous status of the LFS patient, showed the absence of a dominant negative effect. To further confirm our data, we performed functional studies on the variant p.E224A in the colon cancer line HCT116^TP53-/-^, including reporter and clonogenic assays, as well as endogenous P53 target expression analysis; the results confirmed the wild type-like behavior of the mutant P53 protein. Based on these results we assigned to the variant the BS3 criterion (but weighted as supporting toward benign classification), which again was not sufficient for a clear clinical interpretation of the identified *TP53* germline variant c.671A>C (p.E224A) (*i.e.*, still VUS) (**Supplementary Table 2, version 2.1**).

As a matter of fact, the identification of a *TP53* VUS may pose challenges in the genetic counselling and clinical management of patients and their families. VUSs in *TP53* are not diagnostic of LFS; thus, their analysis in family members is not recommended, precluding the chance of addressing other possible carriers for optimal LFS surveillance. However, an accurate revision of literature data, along with *in silico* splicing predictions of the c.671A>C *TP53* variant, prompted us to further investigate its functional impact. A recent large-scale experiment introduced 9,225 *TP53* variants into cancer cells utilizing CRISPR-based technology [19]. Interestingly, the most significant discrepancy compared to previous cDNA-based studies was observed in poorly conserved residues near exon boundaries of the *TP53* gene (such as G187, E224, V225, and S261), highlighting a potential critical role of the splicing in impairing the function of *TP53* missense variants. Specifically, *TP53* variants at codon 224, including synonymous changes (*i.e.*, E224D and E224E), caused reduced mRNA levels and loss of function. Moreover, a different study through *in vitro* experiments with mouse embryonic fibroblasts and human cancer cells, as well as *in vivo* studies with genetically engineered mice, demonstrated that the P53 variant encoding p.E221D (*i.e.*, the mouse equivalent of P53 p.E224D) impairs tumor suppression by causing mis-splicing of the *TP53* mRNA transcript, leading to loss of P53 function [39].

To align c.671A>C (p.E224A) *TP53* variant classification with these data, we developed a splicing functional assay based on a minigene construct in different cell lines, that showed a complete exclusion of *TP53* exon 6 from the transcript of the variant. This splicing alteration was supposed to cause a frameshift in the coding sequence, introducing an early stop codon, likely targeting the resulting *TP53* transcript to NMD.

Therefore, we postulated that the variant could behave as a null allele, thus contributing to the clinical phenotype of the proband; this hypothesis was confirmed by the loss of heterozygosity at the c.671 nucleotide position of *TP53* mRNA obtained from the whole blood of the proband’s parent. The present data are in accordance with the findings of a very recent study, published while our manuscript was in preparation, in which a list of *TP53* variants, including the c.671A>C (p.E224A), has been prioritized based on bioinformatic predictions, confirming the splicing impairment [40].

Finally, these experimental results allowed us to reclassify the variant from VUS as likely pathogenic by applying the full strength weighted PVS1 criterion (*i.e*., generation of aberrant transcripts through mRNA analysis by the variant) in combination with the supporting PM2 criterion (**Supplementary Table2, Version 2.2.0**).

Therefore, the damaging effect of the c.671A>C (p.E224A) *TP53* variant we demonstrated was consistent with the family history, characterized by multiple or bilateral cancers, diagnosed at a young age, and multiple primary tumors in the same individuals. The re-classification of this *TP53* variant proved to be crucial not only for understanding the molecular basis of the child’s cancer but also for other family members who are potential carriers and have an increased risk of developing cancers, thus enabling preventive protocols and follow-up. Indeed, the presence of the c.671A>C (p.E224A) variant was excluded from the proband’s sibling; at the same time, the presence of the variant in the unaffected parent raised the question of an incomplete penetrance of the mutation, leading to the planning of a surveillance strategy for LFS. The clinical management of the family significantly improved as the other carriers now can undergo LFS surveillance.

Surprisingly, exon 6 skipping was also detected using the wild type splicing construct, indicating a weak native donor site and the existence of an alternative RNA processing. A *TP53* transcript lacking exon 6 (NR_176236.1) is annotated as non-coding, with unknown expression and function. In control lymphoblasts, we did not observe exon 6 skipping, even after NMD inhibition (data not shown); these discrepancies may reflect cell-type-specific and condition-dependent splicing regulation. Interestingly, an exon 6–skipped *TP53* isoform was previously detected by RT-PCR in CLL patients, but not in healthy donors; this variant encodes a truncated, transactivation-deficient P53 protein due to a premature stop codon, whose stable expression in H1299 cells enhances growth, suggesting a potential oncogenic role [41,42].

In conclusion, the present work highlights the importance of comprehensive functional analysis in understanding the complex effects of *TP53* variants; indeed, here, we demonstrated that the P53 dysfunction of c.671A>C *TP53* germline missense variant is due to an impairment in the transcript splicing rather than an altered functional activity of the resulting P53 protein p.E224A. Moreover, we also showed that other *TP53* variants at codon 224 might affect P53 functionality through the same mechanism that involves the splicing. Although more than 200 common missense and null *TP53* mutations are well established as disease-causing in LFS, the understanding of the pathogenicity of rare and under-explored *TP53* variants, including splice and small in-frame deletions, is still developing [43,44]. Thus, while the rarity of specific *TP53* variants poses challenges, this study underscores the importance of conducting comprehensive functional studies to accurately assess pathogenicity, thereby supporting genetic counseling and personalized care for individuals with LFS and their at-risk relatives.

## Author Contributions

All authors have seen and approved the manuscript. The manuscript hasn’t been accepted or published elsewhere.

**Conceptualization:** PMo,VC, VG

**Data Curation:** PMo, PMe, RB, VC, FR, FF

**Investigation:** IV, SC, DR, FR, DS, RB, PDM, VG, GC, LM, MO, AG, CM, PH, YC, AB, FF, PMe, PMo, PMa, VC

**Methodology:** IV, SC, RB, SB, PMo, PMe, DS, DR, FR, EM

**Supervision:** PMo, PMe, RB, VC

**Writing – Original Draft Preparation:** PMo, FR, PDM, IV, SC, RB, DR, DS

**Writing – Review & Editing:** PMo, VC, FR, PMe

## Acknowledgements

This work was partially funded by the Italian Ministry of Health (5×1000 funds 2020 to P.Menichini and P.Monti; Current Research 2022-24 to P. Menichini); AIRC IG Grant ID 27852 to P. Malatesta; E.T.S. Engineering Ship Technology Pte Ltd. The authors acknowledge the contributions of ERN ITHACA in improving clinical practice across the EU. We would like to sincerely thank the technical support team at BMR Genomics (Padua, Italy) for their invaluable assistance and expertise throughout this study.

## Data Availability Statement

Data sources and handling of the publicly available datasets used in this study are described in the Materials and Methods. Further details and other data that support the findings of this study are available from the corresponding authors upon request.

## Ethics statement

This study was conducted in accordance with the Declaration of Helsinki, and Ethics approval was obtained from the medical ethics committee established by Gaslini Children’s Hospital (Comitato Etico Territoriale della Regione Liguria, approval code: CER Liguria 399/2021). Written informed consent was obtained from parents for clinical testing and publication of genetic and clinical data.

## References

1. Lane DP, Crawford LV. T antigen is bound to a host protein in SV40-transformed cells. Nature. 1979;278: 261–263. doi:10.1038/278261a0

2. Linzer DI, Maltzman W, Levine AJ. The SV40 A gene product is required for the production of a 54,000 MW cellular tumor antigen. Virology. 1979;98: 308–318. doi:10.1016/0042-6822(79)90554-3

3. Lane DP. Cancer. p53, guardian of the genome. Nature. 1992;358: 15–16. doi:10.1038/358015a0

4. Levine AJ, Oren M. The first 30 years of p53: growing ever more complex. Nat Rev Cancer. 2009;9: 749–758. doi:10.1038/nrc2723

5. Liu Y, Su Z, Tavana O, Gu W. Understanding the complexity of p53 in a new era of tumor suppression. Cancer Cell. 2024;42: 946–967. doi:10.1016/j.ccell.2024.04.009

6. Li FP, Fraumeni JF, Mulvihill JJ, Blattner WA, Dreyfus MG, Tucker MA, et al. A cancer family syndrome in twenty-four kindreds. Cancer Res. 1988;48: 5358–5362.

7. Hosseini M-S. Current insights and future directions of Li-Fraumeni syndrome. Discov Oncol. 2024;15: 561. doi:10.1007/s12672-024-01435-w

8. Kumamoto T, Yamazaki F, Nakano Y, Tamura C, Tashiro S, Hattori H, et al. Medical guidelines for Li-Fraumeni syndrome 2019, version 1.1. Int J Clin Oncol. 2021;26: 2161–2178. doi:10.1007/s10147-021-02011-w

9. Flaman JM, Robert V, Lenglet S, Moreau V, Iggo R, Frebourg T. Identification of human p53 mutations with differential effects on the bax and p21 promoters using functional assays in yeast. Oncogene. 1998;16: 1369–1372. doi:10.1038/sj.onc.1201889

10. Di Como CJ, Prives C. Human tumor-derived p53 proteins exhibit binding site selectivity and temperature sensitivity for transactivation in a yeast-based assay. Oncogene. 1998;16: 2527– 2539. doi:10.1038/sj.onc.1202041

11. Campomenosi P, Monti P, Aprile A, Abbondandolo A, Frebourg T, Gold B, et al. p53 mutants can often transactivate promoters containing a p21 but not Bax or PIG3 responsive elements. Oncogene. 2001;20: 3573–3579. doi:10.1038/sj.onc.1204468

12. Inga A, Monti P, Fronza G, Darden T, Resnick MA. p53 mutants exhibiting enhanced transcriptional activation and altered promoter selectivity are revealed using a sensitive, yeast-based functional assay. Oncogene. 2001;20: 501–513. doi:10.1038/sj.onc.1204116

13. Kato S, Han S-Y, Liu W, Otsuka K, Shibata H, Kanamaru R, et al. Understanding the function-structure and function-mutation relationships of p53 tumor suppressor protein by high-resolution missense mutation analysis. Proc Natl Acad Sci U S A. 2003;100: 8424–8429. doi:10.1073/pnas.1431692100

14. Bisio A, Ciribilli Y, Fronza G, Inga A, Monti P. TP53 mutants in the tower of babel of cancer progression. Hum Mutat. 2014;35: 689–701. doi:10.1002/humu.22514

15. Stein Y, Aloni-Grinstein R, Rotter V. Mutant p53 oncogenicity: dominant-negative or gain-of-function? Carcinogenesis. 2020;41: 1635–1647. doi:10.1093/carcin/bgaa117

16. Boettcher S, Miller PG, Sharma R, McConkey M, Leventhal M, Krivtsov AV, et al. A dominant-negative effect drives selection of TP53 missense mutations in myeloid malignancies. Science. 2019;365: 599–604. doi:10.1126/science.aax3649

17. Giacomelli AO, Yang X, Lintner RE, McFarland JM, Duby M, Kim J, et al. Mutational processes shape the landscape of TP53 mutations in human cancer. Nat Genet. 2018;50: 1381– 1387. doi:10.1038/s41588-018-0204-y

18. Kotler E, Shani O, Goldfeld G, Lotan-Pompan M, Tarcic O, Gershoni A, et al. A Systematic p53 Mutation Library Links Differential Functional Impact to Cancer Mutation Pattern and Evolutionary Conservation. Mol Cell. 2018;71: 178–190.e8. doi:10.1016/j.molcel.2018.06.012

19. Funk JS, Klimovich M, Drangenstein D, Pielhoop O, Hunold P, Borowek A, et al. Deep CRISPR mutagenesis characterizes the functional diversity of TP53 mutations. Nat Genet. 2025;57: 140–153. doi:10.1038/s41588-024-02039-4

20. Capra V, Consales A, Nozza P, Monti P, Inga A, Fronza G. Identification of a novel TP53 germline mutation in a large Italian Li-Fraumeni syndrome Family. Pediatr Blood Cancer. 2009;52: 303–304. doi:10.1002/pbc.21796

21. Monti P, Perfumo C, Bisio A, Ciribilli Y, Menichini P, Russo D, et al. Dominant-negative features of mutant TP53 in germline carriers have limited impact on cancer outcomes. Mol Cancer Res. 2011;9: 271–279. doi:10.1158/1541-7786.MCR-10-0496

22. Monti P, Ciribilli Y, Jordan J, Menichini P, Umbach DM, Resnick MA, et al. Transcriptional functionality of germ line p53 mutants influences cancer phenotype. Clin Cancer Res. 2007;13: 3789–3795. doi:10.1158/1078-0432.CCR-06-2545

23. Penkert J, Strüwe FJ, Dutzmann CM, Doergeloh BB, Montellier E, Freycon C, et al. Genotype-phenotype associations within the Li-Fraumeni spectrum: a report from the German Registry. J Hematol Oncol. 2022;15: 107. doi:10.1186/s13045-022-01332-1

24. Montellier E, Lemonnier N, Penkert J, Freycon C, Blanchet S, Amadou A, et al. Clustering of TP53 variants into functional classes correlates with cancer risk and identifies different phenotypes of Li-Fraumeni syndrome. iScience. 2024;27: 111296. doi:10.1016/j.isci.2024.111296

25. Frebourg T, Bajalica Lagercrantz S, Oliveira C, Magenheim R, Evans DG, European Reference Network GENTURIS. Guidelines for the Li-Fraumeni and heritable TP53-related cancer syndromes. Eur J Hum Genet. 2020;28: 1379–1386. doi:10.1038/s41431-020-0638-4

26. Fortuno C, Pesaran T, Mester J, Dolinsky J, Yussuf A, McGoldrick K, et al. Genotype-phenotype correlations among TP53 carriers: Literature review and analysis of probands undergoing multi-gene panel testing and single-gene testing. Cancer Genet. 2020;248–249: 11–17. doi:10.1016/j.cancergen.2020.09.002

27. de Andrade KC, Khincha PP, Hatton JN, Frone MN, Wegman-Ostrosky T, Mai PL, et al. Cancer incidence, patterns, and genotype-phenotype associations in individuals with pathogenic or likely pathogenic germline TP53 variants: an observational cohort study. Lancet Oncol. 2021;22: 1787–1798. doi:10.1016/S1470-2045(21)00580-5

28. Müntnich LJ, Dutzmann CM, Großhennig A, Härter V, Keymling M, Mastronuzzi A, et al. Cancer risk in carriers of TP53 germline variants grouped into different functional categories. JNCI Cancer Spectr. 2025;9: pkaf008. doi:10.1093/jncics/pkaf008

29. Wang K, Li M, Hakonarson H. ANNOVAR: functional annotation of genetic variants from high-throughput sequencing data. Nucleic Acids Res. 2010;38: e164. doi:10.1093/nar/gkq603

30. Fortuno C, Lee K, Olivier M, Pesaran T, Mai PL, de Andrade KC, et al. Specifications of the ACMG/AMP variant interpretation guidelines for germline TP53 variants. Hum Mutat. 2021;42: 223–236. doi:10.1002/humu.24152

31. Inga A, Storici F, Darden TA, Resnick MA. Differential transactivation by the p53 transcription factor is highly dependent on p53 level and promoter target sequence. Mol Cell Biol. 2002;22: 8612–8625. doi:10.1128/MCB.22.24.8612-8625.2002

32. Burn TC, Connors TD, Klinger KW, Landes GM. Increased exon-trapping efficiency through modifications to the pSPL3 splicing vector. Gene. 1995;161: 183–187. doi:10.1016/0378-1119(95)00223-s

33. Flaman JM, Frebourg T, Moreau V, Charbonnier F, Martin C, Chappuis P, et al. A simple p53 functional assay for screening cell lines, blood, and tumors. Proc Natl Acad Sci U S A. 1995;92: 3963–3967. doi:10.1073/pnas.92.9.3963

34. Inga A, Iannone R, Monti P, Molina F, Bolognesi M, Abbondandolo A, et al. Determining mutational fingerprints at the human p53 locus with a yeast functional assay: a new tool for molecular epidemiology. Oncogene. 1997;14: 1307–1313. doi:10.1038/sj.onc.1200952

35. Garrido-Martín D, Palumbo E, Guigó R, Breschi A. ggsashimi: Sashimi plot revised for browser- and annotation-independent splicing visualization. PLoS Comput Biol. 2018;14: e1006360. doi:10.1371/journal.pcbi.1006360

36. Rocca V, Blandino G, D’Antona L, Iuliano R, Di Agostino S. Li-Fraumeni Syndrome: Mutation of TP53 Is a Biomarker of Hereditary Predisposition to Tumor: New Insights and Advances in the Treatment. Cancers (Basel). 2022;14: 3664. doi:10.3390/cancers14153664

37. Bougeard G, Renaux-Petel M, Flaman J-M, Charbonnier C, Fermey P, Belotti M, et al. Revisiting Li-Fraumeni Syndrome From TP53 Mutation Carriers. J Clin Oncol. 2015;33: 2345– 2352. doi:10.1200/JCO.2014.59.5728

38. Kron A, Alidousty C, Scheffler M, Merkelbach-Bruse S, Seidel D, Riedel R, et al. Impact of TP53 mutation status on systemic treatment outcome in ALK-rearranged non-small-cell lung cancer. Ann Oncol. 2018;29: 2068–2075. doi:10.1093/annonc/mdy333

39. Lock IC, Leisenring NH, Floyd W, Xu ES, Luo L, Ma Y, et al. Mis-splicing Drives Loss of Function of p53E224D Point Mutation. bioRxiv. 2023; 2023.08.01.551439. doi:10.1101/2023.08.01.551439

40. Fortuno C, Llinares-Burguet I, Canson DM, de la Hoya M, Bueno-Martínez E, Sanoguera-Miralles L, et al. Exploring the role of splicing in TP53 variant pathogenicity through predictions and minigene assays. Hum Genomics. 2025;19: 2. doi:10.1186/s40246-024-00714-5

41. Pekova S, Cmejla R, Smolej L, Kozak T, Spacek M, Prucha M. Identification of a novel, transactivation-defective splicing variant of p53 gene in patients with chronic lymphocytic leukemia. Leuk Res. 2008;32: 395–400. doi:10.1016/j.leukres.2007.06.022

42. Pekova S, Mazal O, Cmejla R, Hardekopf DW, Plachy R, Zejskova L, et al. A comprehensive study of TP53 mutations in chronic lymphocytic leukemia: Analysis of 1287 diagnostic and 1148 follow-up CLL samples. Leuk Res. 2011;35: 889–898. doi:10.1016/j.leukres.2010.12.016

43. Louis J, Rolain M, Levacher C, Baudry K, Pujol P, Ruminy P, et al. Li-Fraumeni syndrome: a germline TP53 splice variant reveals a novel physiological alternative transcript. J Med Genet. 2025;62: 160–168. doi:10.1136/jmg-2024-110449

44. Quinn EA, Maciaszek JL, Pinto EM, Phillips AH, Berdy D, Khandwala M, et al. From uncertainty to pathogenicity: clinical and functional interrogation of a rare TP53 in-frame deletion. Cold Spring Harb Mol Case Stud. 2019;5: a003921. doi:10.1101/mcs.a003921

