## Supplementary Materials for "Missense but mis-spliced: germline *TP53* variant c.671A>C (p.E224A) and the path from uncertainty to pathogenicity"

**Supplementary Material**

**Construction of yeast and mammalian vectors expressing the P53 variant p.E224A.** For the generation of yeast plasmids expressing the P53 protein p.E224A, the pLS76 plasmid was used as source for the amplification of WT *TP53* coding sequence [1]. Firstly, a pair of complementary 30-mer oligonucleotides, with the mutated base adjacent to the central position of the sequence, were used in two separate PCR reactions as forward (5’-TATGAGCCGCCT**GCG**GTTGGCTCTGACTGT-3’) and reverse (5’-ACAGTCAGAGCCAAC**CGC**AGGCGGCTCATA-3’) primers (ThermoFisher Scientific by BMR Genomics, Padua, Italy) and paired with P4 and P3 primers [1], respectively. Pfu DNA Polymerase was used for PCR reactions (Biotech Rabbit, Germany). Unpurified aliquots of PCR products were co-transformed in the yIG397 yeast strain together with a HindIII/StuI (New England Biolabs, USA) double digested pRDI-22 plasmid (pLS-based, ADH1 constitutive promoter, LEU2 selection marker); by exploiting the specific sequence homology of the fragments (Gap Repair Assay), the plasmid was re-sealed together with the PCR products by recombination [1]. Plasmid DNA (pLS-p.E224A) was recovered from yeast colonies, expanded in *E.coli* competent cells, extracted (GeneElute, Plasmid Miniprep Kit, Sigma-Aldrich, USA) and checked by Sanger sequencing (BMR Genomics) for the presence of the specific *TP53* variant (c.671A>C). The pTSG-based vector (GAL1,10 inducible promoter, TRP1 selection marker) expressing the P53 protein p.E224A (pTSG-p.E224A) was created from the available pLS-p.E224A vector by SgraI/StuI digestion and subsequent ligation of the fragment with the nucleotide change into an identical double-digested pTSG backbone; as previously described, after the expansion in bacterial competent cells and extraction, the presence of the specific *TP53* variant was confirmed by Sanger sequencing [2]. The mammalian pCI-neo vector expressing the protein p. E224A was obtained from the yeast pTSG-based vectors by XhoI/NotI double digestion and subsequent ligation into an identical double-digested pCI-neo plasmid.

**Yeast reporter assay.** pLS89, pTSG-P53 and pTSG-p.R175H vectors (inducible GAL1,10 promoter; TRP1 selection marker) were already available as well as pLS76 (wild type P53) and pLS-p.R175H vectors (constitutive ADH1 promoter; LEU2 selection marker [2]; plasmids pLS89 and pLS76 encodes for the wild type P53 coding sequence. pRS314 (TRP1) and pRS315 (LEU2) were used as empty vectors. The transactivation activity of P53 proteins (wild type: WT, p.R175H and p.E.224A) was evaluated by transforming the yLFM-P21-5′, yLFM-MDM2P2C, yLFM-BAX A + B and yLFM-PUMA yeast strains with the corresponding pTSG-based vectors. Transformants were selected on minimal plates lacking tryptophan but containing 200 mg/L adenine; after 3 days of growth, the transformants were streaked onto the same type of plate and allowed to grow for 1 day. Yeast cells were then resuspended in selective medium containing raffinose plus 0.128% galactose and grown for 8 hours at 30°C or 37°C in a transparent 96-well plate. The dominant negative activity of P53 proteins (WT, p.R175H and p.E224A) was evaluated similarly by i) co-transforming the same yeast reporter strains with pLS89 plus empty pRS315 (*i.e.*, single WT P53 protein expression) or pLS89 plus pLS-based vectors (pLS76, pLS-p.R175H or pLS-P.E224A: i.e., co-expression of WT and mutant P53 protein), ii) by selecting on the same plates as previously described but in the absence of tryptophan and leucine and iii) by growing yeast transformants in selective medium as above (30°C) plus 0.016% galactose. The reporter assays were performed following the miniaturized protocol we previously developed [3]. Briefly, after 8 hours of growth (30°C or 37°C) in presence of galactose, OD (600 nm) was directly measured in the transparent 96-well plate; then, 20 μl of the cells suspension was transferred into a white 96-well plate and mixed with an equal volume of PLB buffer 2X (Promega, Milan, Italy) to obtain the lysis of yeast cells; after 15 minutes of shaking at room temperature, 20 μl of firefly luciferase substrate (Bright Glo, Promega) was added. Luciferase activity was measured using a multilabel plate reader (Mithras LB940, Berthold Technologies, Germany) and normalized to OD 600 nm.

**Mammalian reporter assay.** The pGL3-P21, pGL3-MDM2, and pGL3-BAX reporter vectors were exploited along with the pRL-SV40 plasmid for the normalization of the transfection efficiency [4]. HCT116*^TP53^*^-/-^ cells were seeded in 24-well plates (1.5 x 10^5^ cells/well) and transfected 24 hours later using Mirus Bio^TM^ TransIT^TM^-LT1 Transfection Reagent (reagent/DNA ratio 3:1) (Mirus Bio LLC, Madison, WI, USA). Two hundred fifty ng of the pGL3-based reporter plasmid, 200 ng of the expression or empty vector and 50 ng of the pRL-SV40 plasmid were mixed in a serum free medium and used for every single transfection; after 24 hours cells were harvested and lysed in 100 μl of 2x Passive Lysis Buffer (PLB, Promega) by shaking for 15 min. Twenty μl of firefly luciferase substrate (Dual Luciferase Reporter Assay, Promega) were added to 20 μl of the cell lysate in a white 96-well plate; pRL-SV40 activity was normalized by adding 20 μl of Stop and Glo solution; luciferase activity was measured as previously described.

**Yeast and mammalian protein extracts**. Yeast pellets were resuspended in an equal volume of water and 0.2 M NaOH (i.e., 100 μL), incubated for 5 minutes at room temperature and centrifuged. The cell pellets were resuspended in 50 μL of SDS sample buffer (Tris HCl 0.06 M pH 6.8, 5% glycerol, 2% SDS, 4% β-mercaptoethanol, 0.0025% bromophenol blue) and boiled at 95°C for 3 minutes; after centrifugation, the supernatants were directly used for western blot analysis. Mammalian cells were harvested by centrifugation at 13,200 rpm for 5 minutes and washed with PBS; lysis was achieved by incubating the cells in lysis buffer (50 mM Tris/HCl pH 7.5, 150 mM NaCl, 1% NP-40, 10% glycerol, 10 mM EDTA, 1 mM DTT) along a protease inhibitors cocktail (Roche, Basel, Switzerland) at 4°C for 30 minutes. After centrifugation, supernatants were collected and protein concentration was determined by BCA assay (ThermoScientific).

**Western Blot.** Protein samples (10-20 μL for yeast extracts; 10-20 μg for mammalian extracts), along with a molecular weight marker (Bio-Rad, Milan, Italy), were separated on 4-15% Mini-Protean TGX Precast gels (Bio-Rad), subjected to electrophoresis at 180 V for 35 minutes, and then transferred to nitrocellulose membranes by Trans-Blot Turbo Blotting System (Bio-Rad). Membranes were blocked with 2% non-fat dry milk in 0.1% Tween-20 in PBS for 1 hour and incubated at room temperature or overnight at 4°C with the specific primary antibodies. The following primary antibodies were used: anti-P53 (clone DO-1, Santa Cruz Biotechnology, Dallas, TX, USA), anti-P21 Waf1/Cip1 (DCS60 Cell Signaling Technology, Danvers, MA, USA), anti-PGK1 (Novex, Thermo Fisher Scientific, Waltham, MA, USA) and anti-human β-Actin (clone AC-74, Sigma-Aldrich, St. Louis, MO, USA), diluted in PBS-T with 1% milk. The appropriate anti-mouse IgG-horseradish peroxidase-conjugated secondary antibody (A9044, Sigma-Aldrich) was used. The detection was performed with ECL FAST PICO (ECL-1002, Immunological Sciences, Rome, Italy), and chemiluminescence was analyzed by Alliance LD, UVITEC Cambridge (Cambridge, U.K.).

**Tables**

| **Supplementary Table 1. *TP53* variant c.671A>C (p.E224A) evidences.** | |
| --- | --- |
| **Functional coding *in silico* prediction** | |
| **Predictor** | **Score** |
| **Align GVGD** | **C35** |
| Grantham variation GV | 29.27 |
| Grantham deviation GD | 89.55 |
| **CADD-Splice scaled score** | **34** |
| **PolyPhen-2** | **Possibly damaging** |
| HumDiv probability of being damaging | 0.606 |
| HumVar probability of being damaging | 0.831 |
| **SIFT** | **Deleterious** |
| SIFT weight | 0.03 |
| SIFT median | 3.53 |
| **MutationTaster** | **Deleterious** |
| Tree vote | 77\|23 [del\|benign] |
| **REVEL** | **Deleterious** |
| Score | 0.825 |
| **MAPP** | **Bad** |
| MAPP p-value | 0.00009112 |
| MAPP p-value median | 0.0002207 |
| **BayesDel** | **Deleterious** |
| noAF meta-score | 0.165391 |
| **Splice altering *in silico* prediction** | |
| **Predictor** | **Score** |
| **SpliceAI** | **Splice-Altering: strong** |
| Donor loss | 0.7673 |
| Donor gain | 0.1941 |
| Acceptor loss | 0.0000 |
| Acceptor gain | 0.0004 |
| **MaxEntScan** | **-100%** |
| **SpliceSiteFinder** | **-12.4%** |
| **GeneSplicer** | **-100%** |
| **NNSPLICE** | -93.2% |
| **dbscSNV** | **Deleterious** |
| Ada score | 1 |
| RF score | 0.97 |
| **Population data** | |
| **gnomAD v2.1.1 (non cancer)** | **Not referenced** |
| PLI | 0.53235 |
| ExAc PLI | 0.91222 |
| Observed/Expected ratio | 0.20 (0.10-0.47) |
| LOEUF percentile | 0.27206 |
| **Databases/Repository** | |
| **ClinVar** | **** Uncertain significance** |
| Variation ID | 428892 |
| Accession | VCV000428892.15 |
| **LOVD** | **Not classified** |
| Effect | ?/. (unknown) |
| Frequency | 1/60,466 cases |

| **Supplementary Table 2. P53 protein p.E224A activity in reporter assays.** | | | | |
| --- | --- | --- | --- | --- |
| **Yeast cells-based assay** | | | | |
| **Transactivation ability (30°C)** | **yLFM-P21-5'** | **yLFM-MDM2P2C** | **yLFM-**  **BAX A+B** | **yLFM-PUMA** |
| WT | 100 ± 8 | 100 ± 3 | 100 ± 7 | 100 ± 6 |
| p.R175H | 0 ± 0 | 1 ± 0 | 1 ± 0 | 1 ± 0 |
| p.E224A | 98 ± 16 | 123 ± 8 | 108 ± 12 | 85 ± 6 |
| **Transactivation ability (37°C)** | **yLFM-P21-5'** | **yLFM-MDM2P2C** | **yLFM-**  **BAX A+B** | **yLFM-PUMA** |
| WT | 100 ± 7 | 100 ± 5 | 100 ± 4 | 100 ± 3 |
| p.R175H | 1 ± 0 | 1 ± 0 | 2 ± 0 | 1 ± 0 |
| p.E224A | 105 ± 15 | 125 ± 8 | 116 ± 10 | 85 ± 8 |
| **Dominant negative ability (30°C)** | **yLFM-P21-5'** | **yLFM-MDM2P2C** | **yLFM-**  **BAX A+B** | **yLFM-PUMA** |
| WT + EV | 100 ± 13 | 100 ± 18 | 100 ± 23 | 100 ± 7 |
| WT + p.R175H | 66 ± 11 | 49 ± 7 | 65 ± 9 | 59 ± 11 |
| WT + p.E224A | 378 ± 32 | 478 ± 41 | 1026 ± 43 | 846 ± 90 |
| WT + WT | 370 ± 43 | 504 ± 44 | 761 ± 165 | 801 ± 48 |
| **Mammalian cells-based assay** | | | | |
| **Transactivation ability** | | **pGL3-**  **P21** | **pGL3-**  **MDM2** | **pGL3-**  **BAX** |
| WT |  | 100 ± 18 | 100 ± 16 | 100 ± 13 |
| p.R175H |  | 11 ± 2 | 9 ± 1 | 1 ± 0 |
| p.E224A |  | 123 ± 5 | 128 ± 19 | 110 ± 17 |

| **Supplementary Table 3. *TP53* variants at codon 224 in somatic and germline datasets.** | | | | | | |
| --- | --- | --- | --- | --- | --- | --- |
| **Protein change** | **Nucleotide substitution** | **cDNA position** | **Splice AI (donor loss)** | **Germline frequency (n° families in IARC)** | **COSMIC frequency** | **GnomAD frequency** |
| **p.E224A** | **GAG-->GCG** | **c.671A>C** | **0,77** | **0** | **0** | **NA** |
| p.E224D | GAG-->GAC | c.672G>C | 0,98 | 0 | 17 | NA |
| p.E224D | GAG-->GAT | c.672G>T | 0,97 | 1 | 27 | 1,59055E-06 |
| p.E224E | GAG-->GAA | c.672G>A | 0,90 | 0 | 13 | NA |
| p.E224G | GAG-->GGG | c.671A>G | 0,73 | 0 | 2 | NA |
| p.E224K | GAG-->AAG | c.670G>A | 0,09 | 0 | 8 | NA |
| p.E224Q | GAG-->CAG | c.670G>C | 0,08 | 0 | 1 | 6,84E-07 |
| p.E224V | GAG-->GTG | c.671A>T | 0,85 | 0 | 1 | NA |

| **Supplementary Table 4. Classification of *TP53* variant c.671A>C (p.E224A)** | | | | |
| --- | --- | --- | --- | --- |
| **Rule code** | **Evidence strength** | **Criteria**  **description** | **ClinGen *TP53* guidelines** | |
|  |  |  | **Version 1.2** | **Version 2.2.0** |
| **PM2** | Supporting | Absent in population databases | **✓** | **✓** |
|  |  |  | Not found in gnomAD database | Not found in gnomAD database |
| **PP3** | Supporting | Multiple lines of computational evidence support a deleterious effect on the gene or gene product | **✓** | **X** |
|  |  |  | Concordance of two predictors (A-GVGD C35; BayesDel 0.17) | PP3 should not be used in combination with PVS1 |
| **BS3** | Supporting | Well-established *in vitro* or *in vivo* functional studies show no damaging effect on protein function or splicing | **✓** | **X** |
|  |  |  | Transactivation assays in yeast (IARC classification based on data from Kato et al, 2003) that demonstrate a partially functioning allele (>20% and <=75% activity) **AND** There is a 2nd assay demonstrating retained function (here reported *in vitro* yeast model) | Studies conducted in human cell lines indicate this alteration is proficient at growth suppression and has a dominant negative effect (Kotler et al, 2018; Giacomelli et al, 2018) |
| **PVS1** | Very strong | In a gene where loss of function (LOF) is a known mechanism of disease | **X** | **✓** |
|  |  |  | Not applicable following workflow guidance provided in Tayoun et al, 2018 | Variant inducing aberrant transcripts identified via mRNA assay (minigene splicing assay here reported) following recommendations from Walker et al, 2023 |
| **FINAL CLASSIFICATION** | | | **Uncertain**  **Significance** | **Likely**  **Pathogenic** |

**Figures**

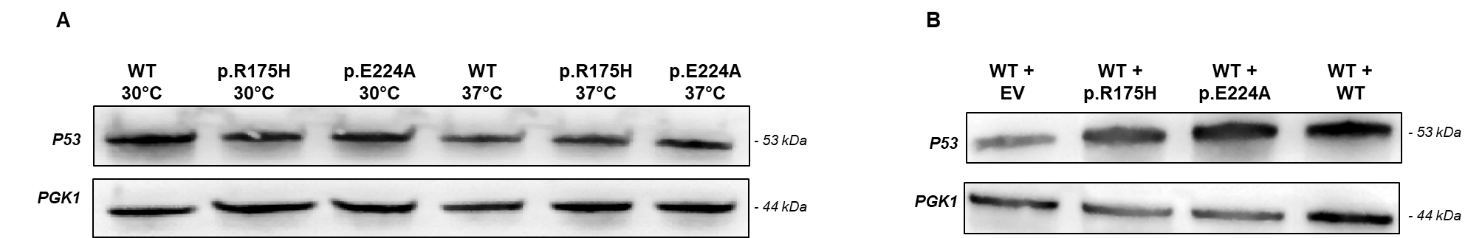

**Supplementary Figure 1**. Western Blot analysis of P53 proteins from the A) transactivation (30°C and 37°C) and B) dominant negative (30°C) reporter experiments in yLFM-PUMA strain. PGK1 protein loading is shown as a housekeeping gene. Western blot analysis was performed in one strain, (*i.e.*, yLFM-PUMA), since all strains are isogenic and differ only for the RE sequence (20 bps) upstream of the luciferase reporter gene.

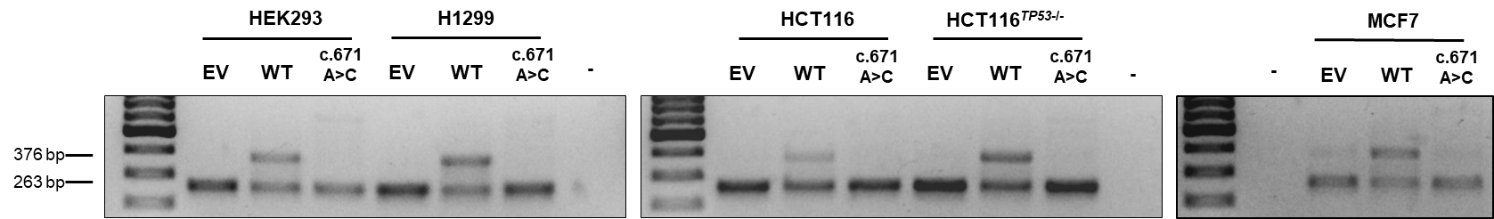

**Supplementary Figure 2.** Wild type and mutant genomic *TP53* fragments (c.671A>C) subcloned into the pSPL3 splicing vector were transfected in HEK293 cells along with the cell lines H1299, HCT116, HCT116*^TP53^*^-/-^and MCF7. PCR products obtained from empty (EV), wild type (WT), mutant vectors and negative control (-) are shown.

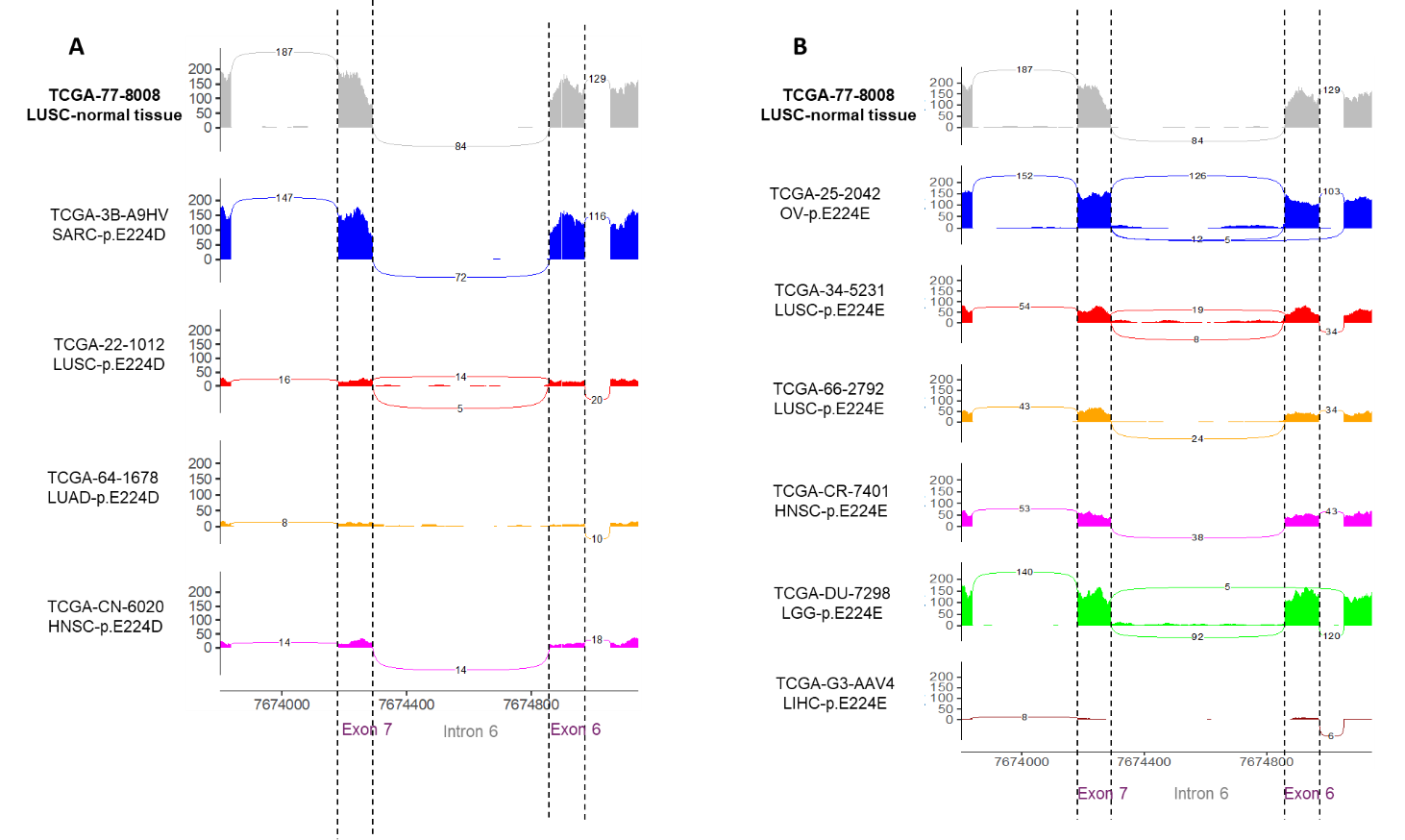

**Supplementary Figure 3**. Sashimi plots showing raw read alignments from TCGA patients with *TP53* variant A) p.E224D: all 4 patients have c.672G>T nucleotide substitution with exception of TCGA-22-1012 (c.672G>C) and B) p.E224E: all 6 patients have c.672G>A nucleotide substitution. RNA splicing patterns in tumor-adjacent normal tissue of a lung cancer patient are shown for comparison. LUSC, lung squamous cell carcinoma; SARC, sarcoma; LUAD, lung adenocarcinoma; HNSC, head and neck squamous cell carcinoma; OV, ovarian cancer; LGG, low grade glioma; LIHC, liver hepatocellular carcinoma.

**References**

1.	Ishioka C, Frebourg T, Yan YX, Vidal M, Friend SH, Schmidt S, et al. Screening patients for heterozygous p53 mutations using a functional assay in yeast. Nat Genet. 1993;5: 124–9. doi:10.1038/ng1093-124

2.	Monti P, Lionetti M, De Luca G, Menichini P, Recchia AG, Matis S, et al. Time to first treatment and P53 dysfunction in chronic lymphocytic leukaemia: results of the O-CLL1 study in early stage patients. Sci Rep. 2020;10: 18427. doi:10.1038/s41598-020-75364-3

3.	Andreotti V, Ciribilli Y, Monti P, Bisio A, Lion M, Jordan J, et al. p53 transactivation and the impact of mutations, cofactors and small molecules using a simplified yeast-based screening system. PLoS One. 2011;6: e20643. doi:10.1371/journal.pone.0020643

4.	Monti P, Russo D, Bocciardi R, Foggetti G, Menichini P, Divizia MT, et al. EEC- and ADULT-associated TP63 mutations exhibit functional heterogeneity toward P63 responsive sequences. Hum Mutat. 2013;34: 894–904. doi:10.1002/humu.22304

**Original Western Blot membranes**

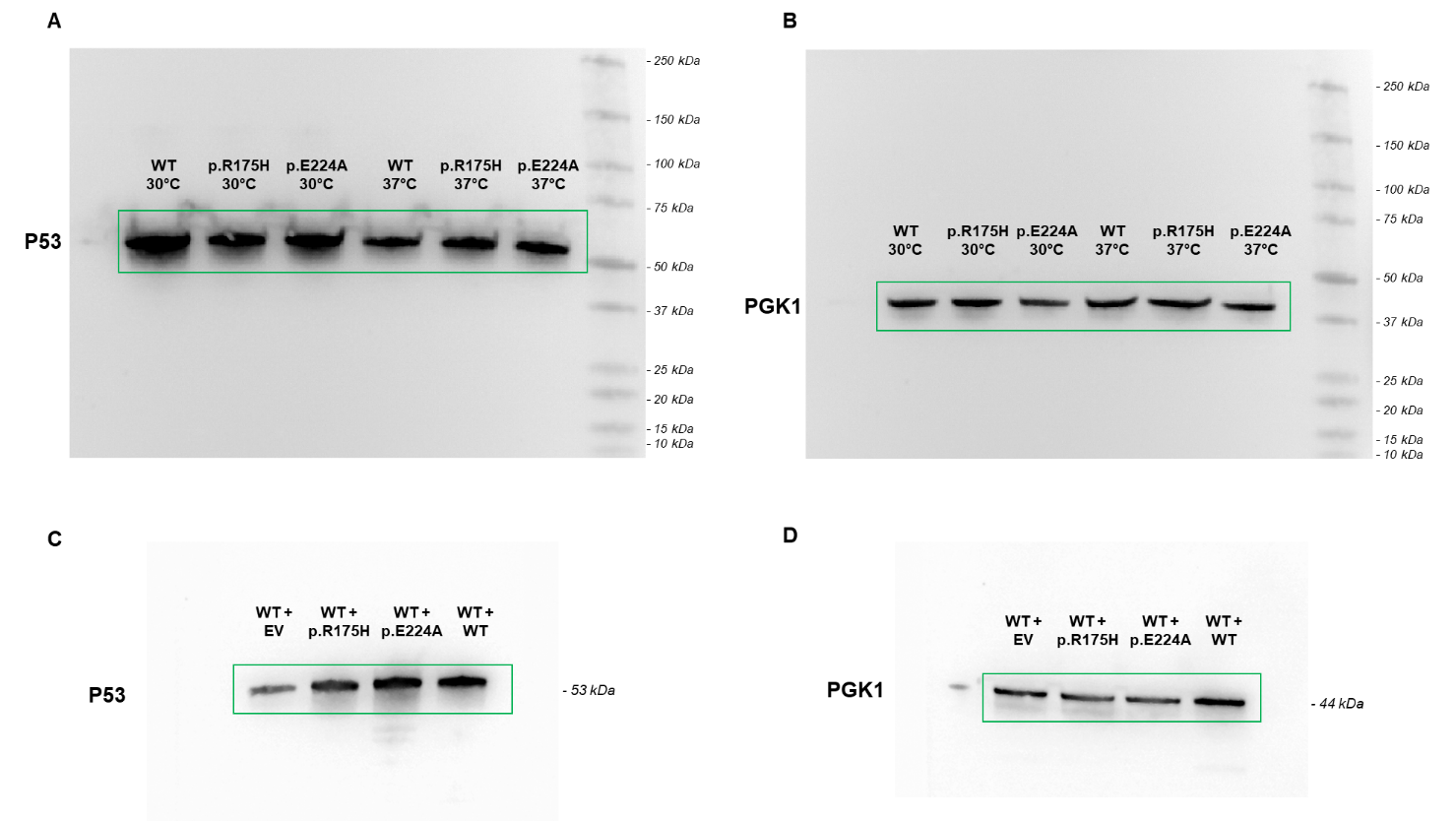

**Full-length blots of proteins illustrated in Supplementary Figure 1**. Yeast extracts from reporter experiments in yLFM-PUMA strain *(i.e.*, transactivation activity at 30°C and 37°C: A and B; dominant negative activity at 30°C: C and D) were loaded on 4–15% Mini Protean TGX precast gels (see M&M). The samples for each reporter assay were run on two separate gels and transferred to two different membranes due to the similar molecular weights of the two target proteins. One membrane was hybridized with anti-P53-DO1 ab and the chemiluminescence was acquired by UVITEC (A and C). The second membrane was hybridized with anti-PGK1 ab and the chemiluminescence was acquired by UVITEC (B and D). The green squares outline the cropped areas are shown in **Supplementary Figure 1**. The images of the membranes are merged with the molecular weight markers.

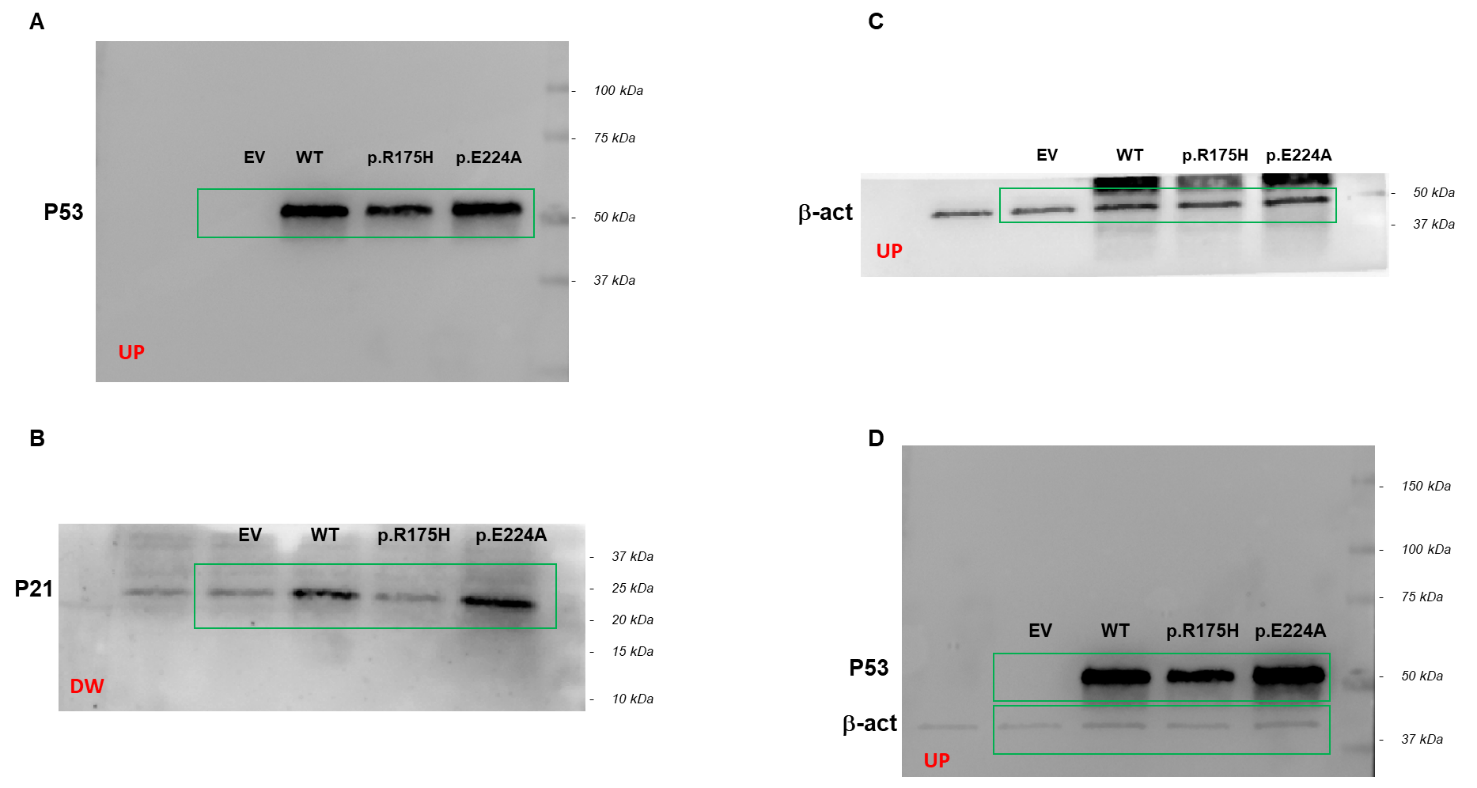

**Full-length blots of proteins illustrated in Figure 2.** Cell extracts from HCT116^TP53-/-^ cells transfected with vectors expressing empty vector (EV), wild-type P53 (WT), p.R175H and p.E224A were loaded on 4-15% Mini Protean TGX precast gels (see M&M). The membrane was cut into two parts, UP and DW, and hybridized with anti-P53-DO1 anti-P21 antibodies, respectively, and the chemiluminescence was acquired by UVITEC (A and B). The same UP membrane was then hybridized with β-actin ab (C) and acquired with UVITEC. The image D shows the full membrane after P53-DO1 and β-actin acquisition. The green squares outline the cropped areas are shown in **Figure 2**. The images of the membranes are merged with the molecular weight markers.
